# Prevalence of anti-SARS-CoV-2 antibodies in people attending the two main Goma markets in the eastern Democratic Republic of Congo

**DOI:** 10.1101/2023.02.17.23286083

**Authors:** Mitangala Ndeba Prudence, Irenge Mwana Wa Bene Léonid, Musubao Tsongo Edgar, Kahindo Mbeva Jean Bosco, Ayonga Ndeba Patrick, Safari Kyembwa Israël, Kubuya Bonane Janvier, Ntabe Namegabe Edmon, Kabangwa Kakongo Senga Raphaël, Mutombo Ndongala Guy, Jérôme Ambroise, Jean-Luc Gala

## Abstract

According to official data, the Democratic Republic of the Congo (DRC) has a low prevalence of the coronavirus disease 19 (COVID-19) pandemic. The goal of this cross-sectional study was to determine the COVID-19 seroprevalence in people attending Goma’s two largest markets, Kituku and Virunga. This study was conducted between September and November 2021, overlapping by one month with another similar study carried out in a slum of Bukavu, and using the same methodology.

COVID-19 unvaccinated participants (n = 796 including 454 vendors and 342 customers, 60% of whom were women) were surveyed. The median age of vendors and customers was 34.2 and 30.1 years, respectively.

The crude and adjusted anti-SARS-CoV-2 antibody seroprevalence rates were 70.2% (95 % CI 66.9-73.4%) and 98.8% (95% CI 94.1-100%), respectively, with no difference between vendors and customers. COVID-19 symptoms were mild or absent in 58.9% and 41.1% of participants with anti-SARS-CoV-2 antibodies respectively. COVID-19 did not require hospitalisation for any of the seropositive participants.

These findings are consistent with those reported in Bukavu. They confirm that SARS-CoV-2 spread without causing severe symptoms in densely populated settlements and markets, and suggest that many COVID-19 cases went unreported. Based on these results, relevance of an untargeted hypothetical vaccination programme in these communities should be questioned.

## Introduction

The coronavirus disease 2019 (COVID-19), a pandemic that swept the world at the end of 2019 is caused by Severe Acute Respiratory Syndrome Coronavirus-2 (SARS-CoV-2), an RNA virus beta-CoV of group 2B virus which was originally described in the Chinese province of Hubei [1]. The pandemic has had a serious impact on global health and the world economy. Two years after the start of the COVID-19 pandemic, and despite alarming predictions about its spread and major impact on the health and mortality of local populations in Africa [2-5], the resilience of sub-Saharan African countries has been much better than expected. This observation is reinforced by the few reports of severe cases and deaths due to COVID-19 in sub-Saharan Africa the last two years [5, 6].

The DRC, which reported its first COVID-19 case on March 10^th^, 2020 [7], is one of the countries that has so far reported a rather low number of COVID-19 cases. As of January 1^st^, 2023, this 90million-person country had reported 95 257 COVID-19 cases and 1460 deaths directly attributed to COVID-19 [8]. The North Kivu province, which borders Uganda and Rwanda, reported its first COVID-19 case on March 30^th^, 2020 [9]. It has since confirmed a total of 11 007 cases, and 603 deaths related to COVID-19 [9] for a population of around 9 million inhabitants [10]. These low figures reported for the North-Kivu province, but also for the DRC, are intriguing given the virtual absence of measures to contain the spread of the SARS-CoV-2 in the country’s cities and towns. In addition, DRC has one of the lowest vaccination rates in the world, with less than 3% of the population fully vaccinated [11] [12]. In the city of Goma, the current vaccination rate is probably one of the lowest ever, with only 0.14% having been completely vaccinated [13].

The country’s capacity to identify the SARS-CoV-2 by real-time PCR is very limited and does not fulfil the World Health Organization guidelines for SARS-CoV-2 screening at the national level [14]. This shortcoming is one of the reasons suggesting some under-reporting of COVID-19 cases and deaths in the DRC. It has also been proposed that the population, as a result of the variety of recurring infectious diseases to which it is chronically exposed, has been able to rapidly acquire effective protective immunity against severe forms of COVID-19, spurred by the high circulation of SARS-CoV-2 [6]. This hypothesis would, at least partially explain the low demand of medical care and COVID-19 related death rate compared to other continents and contribute to the under-reporting of cases. In addition to this hypothesis, the scarcity of epidemiological surveys on the prevalence of COVID-19 in the country drastically hinders assessment of the true scale of the COVID-19 pandemic in the country. It should be noted that serological analyses in this country of 90-million inhabitants are only available in the cities of Kinshasa and Bukavu, and that these seroprevalence data suggest a SARS-CoV-2 circulation, resulting in anti-SARS-CoV-2 antibodies in a significant number of survey participants [15].

Bordered to the north by Lake Kivu, Goma is one of the largest and busiest cities in the DRC. It is an important communication hub between Rwanda, Uganda, Kenya, Kinshasa, Bukavu, and Bujumbura as well as the DRC’s hinterland. Given its location and connections, Goma is a likely hotspot for the rapid expansion of COVID-19 amongst its population and beyond.

The purpose of this survey was to compare the seroprevalence of anti-SARS-CoV-2 antibodies in Bukavu [6] to the seroprevalence in the crowded markets of Kituku and Virunga of Goma city. Bukavu and Goma surveys were conducted three months apart, from June to September 2021 and September to November 2021, respectively. This was done to avoid major differences due to the pandemic’s temporal evolution.

## Material and methods

Between September 1^st,^ 2021, and November 8^th,^ 2021, we conducted a survey to determine the seroprevalence of anti-SARS-CoV-2 antibodies among a sample of vendors, and customers in the Kituku and Virunga markets of Goma, DRC. Goma, a city of about 1 million inhabitants, is located on the northern shore of Lake Kivu, straddling the Rwandan border. It is a major commercial hub in the country’s eastern region and home of several markets, notably the Virunga market (Northeast of Goma) and Kituku market (west of Goma). At midday, the number of people present at Kituku and Virunga markets is around 20 000 in each, with approximatively 4000 vendors in each market.

The sample size of vendors to be enrolled in the study was calculated using the sample size calculator software [16], based on the following parameters: i) a confidence level of 95%, a prevalence (population proportion) of 50%, a margin of error of 5%, and a target population of 30 000. The sample size was augmented by 20% in order to compensate for non-consenting or absent vendors on the day of sampling, which set the number of participants at 456, with a minimum of 380 participants.

### Data collection

A team of 7 members (i.e., outreach members, interviewers, and a laboratory technician) was sent to the two markets to carry out the sampling and collect the questionnaires. Following consent of the market managers, a systematic random sampling of one out of every ten sellers in each row of the market was carried out. The market customers were recruited through convenience sampling, and enrolled in the study at the same time. This sampling entailed enrolling each customer who agreed to participate and was present at a surveyed vendor stand. First, as in a parallel study [6,] selected participants (i.e., vendors and customers) were interviewed to assess their disease knowledge using a questionnaire and the presence of symptoms commonly associated with COVID-19 over the previous six months. Subsequently, fresh finger-prick blood was taken from each participant and placed directly into the well of the QuickZen® COVID-19 IgM/IgG kit (ZenTech, Angleur, Belgium), an immune colloidal gold lateral flow test kit which detects IgM and IgG against SARS-CoV-2 S-RBD (receptor-binding domain of the S protein of SARS-CoV-2). The results were interpreted according to the manufacturer’s recommendations. In a parallel survey carried out in Bukavu in the same time period, 49 pre-COVID-19 serum samples which were collected in the district of Kadutu between April 2004 and May 2005 tested negative with the QuickZen® assay [6]. The same assay was also tested in this study on an additional batch of 32 pre-COVID-19 plasma samples from the Goma population, which were collected in 2019 and stored at the AMI-LABO facility in Goma city.

### Statistical analyses

Statistical analyses were performed using the SPSS statistical package for Windows, version 26.0 (SPSS, Inc., Chicago, IL). The crude seroprevalence was calculated a as the proportion of participants positive for anti-SARS-CoV-2 antibodies. The adjusted seroprevalence was calculated using the standard correction formula published by Sempos and Tian [17]: Adjusted Prevalence = Crude prevalence × specificity - 1)/ sensitivity + specificity - 1

The sensitivity and the specificity of the QuickZen® COVID-19 IgM/IgG kit were previously calculated by Montenisos et al, [18]. Differences in group proportions and categorical variables were assessed using the chi-square test. Odds ratios (ORs) for presence of symptoms associated with COVID-19 in the presence or absence of anti-SARS-CoV-2 antibodies were calculated. A *P* value <0.05 was considered as statistically significant. Adjusted *P* values were computed in R v.4.1.1 with the method of Benjamini Hochberg [19].

### Ethical considerations

The Université Catholique de Bukavu’s Internal Review Board (UCB/CIES/NC/022/2021) reviewed and approved this study. Before enrolment and sample collection by local first-line responders, all participants provided their consent, but given the low level of literacy of market attendees, only verbal consent was requested and recorded. Healthcare workers and physicians signed the following statement: “We have explained the study to the participants and are satisfied that he/she understands and consents to participate in the survey”.

## Results

In total, 796 participants (i.e., 454 vendors and 342 market customers) were included in the survey. Table 1 compares the socio-demographic characteristics of vendors and customers. The median age of vendors was 34.2 and vendors under 40 years old constituted 80.6% of all vendors. Customers were younger, with the median in this group of 30.1 years, and customers under 40 years making 87.7% of all customers interviewed.

**Table 1.** Socio-demographic characteristics and SARS-CoV-2 serology of vendors and customers in two Goma markets

| Variable | Study group |  | P value | SARS-CoV-2 serology |  |  |  |  |  |
| --- | --- | --- | --- | --- | --- | --- | --- | --- | --- |
|  | Vendor<br>n = 454<br>(%) | Customer<br>n = 342<br>(%) |  | Vendors |  |  | Customers |  |  |
|  |  |  |  | Positive | Negative | P value | Positive | Negative | P value |
|  |  |  |  | n = 319<br>(70.3 %) | n = 135<br>(29.7 %) |  | n = 240<br>(70.2 %) | n = 102<br>(29.8 %) |  |
| Goma's markets |  |  |  |  |  |  |  |  |  |
| Kituku | 218 (48) | 163 (47.7) | 0.97 | 152 (69.7) | 66 (30.3) | 0.81 | 112 (68.7) | 51 (31.3) | 0.57 |
| Virunga | 236 (52) | 179 (52.3) |  | 167 (70.8) | 69 (29.2) |  | 128 (71.5) | 51 (28.5) |  |
| Age |  |  |  |  |  |  |  |  |  |
| 18 – 25 yr | 144 (31.7) | 156 (45.6) | <0.01 | 99 (68.8) | 45 (31.2) | 0.81 | 108 (69.2) | 48 (30.8) | 0.94 |
| 26 – 40 yr | 222 (48.9) | 144 (42.1) |  | 156 (70.3) | 66 (29.7) |  | 102 (70.8) | 42 (29.2) |  |
| ≥ 40 yr | 88 (19.4) | 42 (12.3) |  | 64 (72.2) | 24 (27.8) |  | 30 (71.4) | 12 (28.6) |  |
| Sex |  |  |  |  |  |  |  |  |  |
| Female | 296 (65.2) | 179 (52.3) | <0.01 | 210 (70.9) | 86 (29.1) | 0.66 | 127 (70.9) | 52 (29.1) | 0.74 |
| Male | 158 (34.8) | 163 (47.7) |  | 109 (69.0) | 49 (31.0) |  | 113 (69.3) | 50 (30.7) |  |
| Wearing a mask |  |  |  |  |  |  |  |  |  |
| Yes | 80 (17.6) | 85 (24.9) | 0.01 | 55 (68.8) | 25 (31.2) | 0.74 | 59 (69.4) | 26 (30.6) | 0.86 |
| No | 374 (82.4) | 257 (75.1) |  | 264 (70.6) | 110 (29.4) |  | 181 (70.4) | 76 (29.6) |  |
| District |  |  |  |  |  |  |  |  |  |
| Goma | 173 (38.1) | 139 (40.6) | 0.04 | 122 (70.5) | 51 (29.5) | 0.88 | 98 (70.5) | 41 (29.5) | 0.95 |
| Karisimbi | 267 (58.8) | 181 (52.9) |  | 188 (70.4) | 79 (29.6) |  | 126 (69.6) | 55 (30.4) |  |
| Nyiragongo | 14 (3.1) | 22 (6.4) |  | 9 (64.3) | 5 (35.7) |  | 16 (72.7) | 6 (27.3) |  |

In total, 559 participants out of 796 tested positive for anti-SARS-CoV-2 antibodies, resulting in an overall crude seroprevalence of 70.2% (95% CI: 66.9%-73.4%). The adjusted seroprevalence of anti-SARS-CoV-2 antibodies was 98.8% (95% CI 94.1-100%). There was no significant difference between vendors (70.3%, 95 % CI 66.1-74.5%) and customers (70.2%, 95% CI 65.0-75.0%) (*P* value = 0.98). The antibody distribution of anti-SARS-CoV-2 antibodies among positive participants was as follows: IgM 13.4%, IgG 53.4%, IgM and IgG simultaneously 33.2%.

The absence of anti-SARS-CoV2 antibodies in any pre-pandemic sera from Goma and Bukavu [6], or is noteworthy.

There were no significant differences in seroprevalence rates between all groups analysed (vendors vs customers, women vs men, Kituku market participants vs those from the Virunga market). Likewise, no significant differences in seroprevalence were found in age groups.

Both markets were overcrowded, with no social measures implemented in order to prevent the SARS-CoV-2 from spreading among market attendees. Only 17.6% surveyed vendors correctly put on a mask, whereas this rate was significantly higher in customers (24.9%) (*P* value = 0.01).

Symptoms reported by the group of vendors according to their anti-SARS-CoV-2 status are summarized in table 2. Compared with participants who tested negative for the presence of anti-SARS-CoV-2 antibodies (n = 135), those who tested positive (n = 319) were more symptomatic, with odds ratios (OR) consistently higher than 2.0 (adjusted *P* value < 0.05) for each symptom to the exception for ageusia (OR = 1.0 adjusted *P* value = 0.15). 41.2% of participants with anti-SARS-CoV-2 antibodies in the group of vendors did not recall having experienced any COVID-19-related symptoms (OR = 1.0; CI 95% 0.6–1.5%).

**Table 2.**
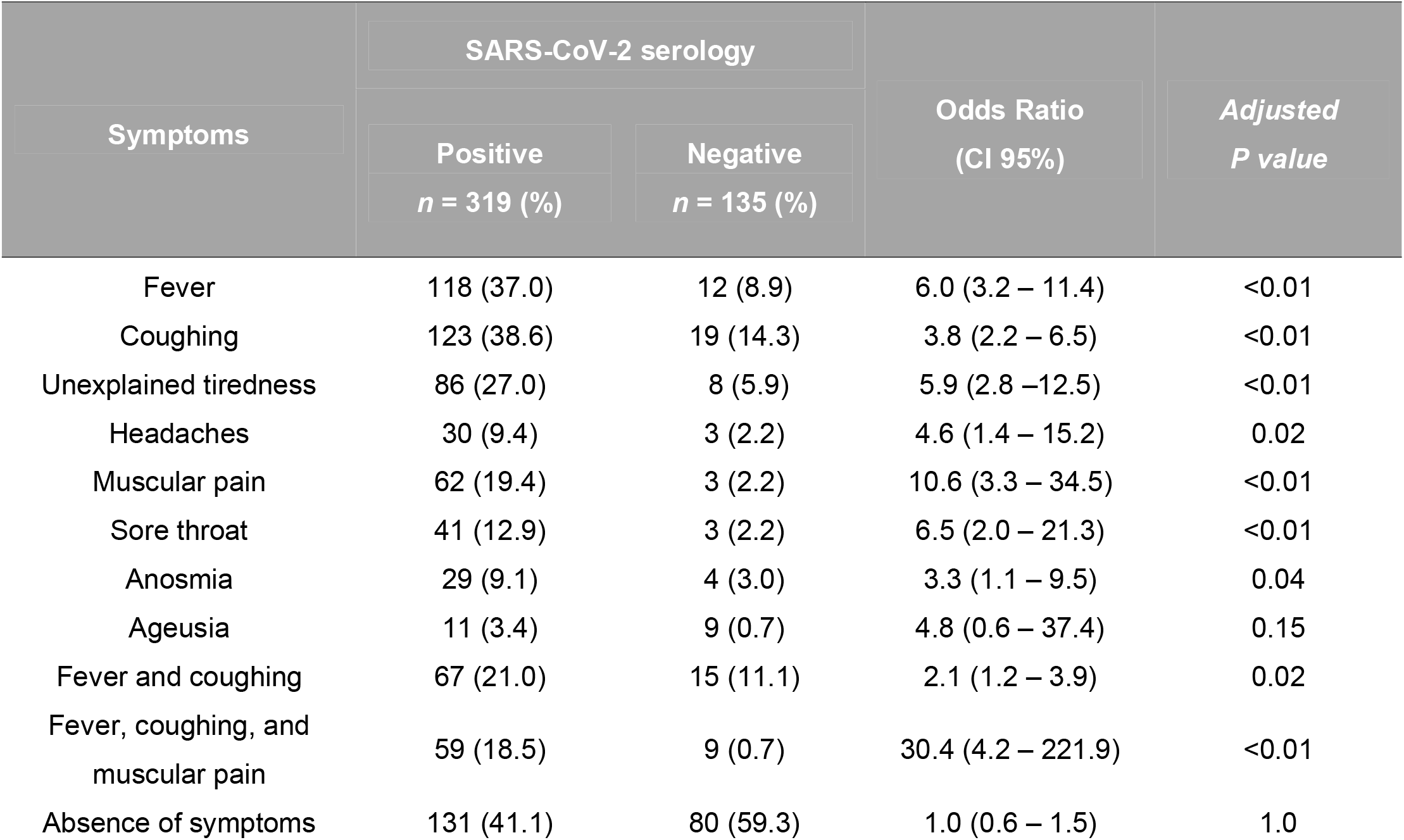
Clinical symptoms and odds ratios among vendors in Kituku and Virunga Goma markets

## Discussion

From September 1^st^ to November 8^th^, 2021, we conducted a seroprevalence survey of anti-SARS-CoV-2 antibodies among vendors and customers in Kituku and Virunga, Goma’s two main markets. The results indicate a seroprevalence rate of 70.3%, and 70.2% among vendors and customers, respectively. The participants’ age or gender did not influence the seroprevalence of anti-SARS-CoV-2 antibodies. The very low proportion of vendors (17.6%) and customers (24.8%) wearing a protective mask correctly is an indication of the weak implementation of preventive measures against the spread of COVID-19, or the lack of thereof in these overcrowded settings. This is worsened by the fact that Goma sellers constantly advertise their wares by shouting, which facilitates virus transmission to market customers via aerosolization [20].

This survey’s seroprevalence rates are among the highest ever documented worldwide in an unvaccinated population. Although these rates cannot be directly extrapolated to the entire city of Goma, the concordance of the seroprevalence rates in the market vendors and customers groups, (even though customers were not sampled randomly) suggests that they are a good indicator of the city’s high seroprevalence rate of anti-SARS-CoV-2 antibodies, hence the potential impact of SARS-CoV-2 transmission in its inhabitants. Moreover, the lack of a guaranteed income, along with frequent power failures, makes it nearly impossible to use refrigerators, and thus to store and preserve perishable goods, compelling most residents to go to markets every day in order to find the products essential to their subsistence. This, combined with the lack of preventive measures as described above, is a major risk factor for SARS-CoV-2 contamination and transmission in their household and neighbourhoods.

The few other serological studies in the DRC have consistently found a high COVID-19 seroprevalence. While a study conducted in Kinshasa at the end of the first wave found had a seroprevalence of 16.6% [21], a recent study conducted in a Bukavu slum and overlapping the current Goma survey by one month, found a much higher COVID-19 seroprevalence with an overall crude seroprevalence rates of 59.7% [6]. The age of the participants in this survey (80% under 40) are comparable with what has been found in the Bukavu survey [6]. Both seroprevalence rates are in line with another serological survey carried out on healthcare personnel of the Panzi hospital in Bukavu with a prevalence of 41.2% of anti-SARS-CoV-2 antibodies [15], and all these studies in Bukavu and Goma confirm the high prevalence of anti-SARS-CoV2 in eastern DRC.

Our study had several limitations. Firstly, specificity and sensitivity of the assay were based on 128 COVID-19 negative sera (i.e. from healthy Belgian volunteers with no recorded contact with SARS-CoV-2), and 72 COVID-19 positive sera from Belgian patients who had displayed of late a positive SARS-CoV-2 signal. In the absence of a multicentric validation of the assay and due to the high number of SARS-CoV-2 variants over the course of this pandemic, the adjustment of African seroprevalence rates based on the sensitivity, and the specificity from the Montesinos study appears to be a potential source of bias. However, all pre-pandemic COVID-19 sera tested negative in Bukavu (n = 49) and Goma (n = 32) confirming the 100% specificity of the test. Second, due to the design of this study and the country’s insufficient testing capacity, our serological data could not be compared to reverse transcription-polymerase chain reaction (RT-PCR) results. Consequently, we could not assess whether SARS-CoV-2 transmission was still active in the market vendors and customers at the time of the survey, and the potential extension thereof. As a result, we were unable to determine the case-to-undetected infection-ratio (CIR) among survey participants due to the lack of RT-PCR results. The DRC health system also lacked trustworthy databases and age-specific population data, making it impossible to calculate the infection fatality rate or adjust seroprevalence for age. The third limitation is that the use of a qualitative anti-SARS-CoV-2 antibody test does not allow the assessment of the amount of antibody required to confer protective immunity against reinfection. The fourth limitation is that we surveyed a cohort of vendors that does not reflect the actual composition of households in Goma. Indeed, children, working-class people, and the elderly are not, a priori, represented in this group where there is an over-representation of women. Finally, customers were selected using a convenience sampling method, which is not a reliable sampling method. Despite these potential biases, the high seroprevalence in this group gives a glimpse of the extent to which SARS-COV-2 might have circulated in the city of Goma, with markets serving as important nodes for its dissemination. Although care should be taken when interpreting the study’s findings and extrapolating them to the entire population of Goma and surroundings, the concordance of the seroprevalence results in both vendors and customers group is indicative of the real serological situation in Goma at the time of the survey. Combined to the data collected at nearly the same period of time in a slum of Bukavu [6], it confirms the widespread SARS-CoV-2 transmission among the population of major cities of eastern DRC in general, and among vendors and customers in Goma’s two largest markets in particular. This may be attributed to a lack of efficient containment measures. Another notable finding of the current study is the absence of severe COVID-19 cases among the participants despite a high rate of antibodies positivity and a total lack of vaccination in the survey participants (i.e., market vendors and customers). The low morbidity rate in surveyed participants is consistent with the recent data reported in Bukavu [6], and in sub-Saharan African countries [22, 23]. While the relatively young age of the surveyed cohort contributes to the low COVID-19 morbidity, these results support the hypothesis of a genuine immunity potential in the population of these crowed markets due to a continuous stimulation by recurrent infectious diseases, among which the SARS-CoV-2 [6]. This non-vaccination related acquired protection, combined with a very low capacity to detect the presence of the SARS-CoV-2 both in local communities and large cities, may contribute to the global under-estimation of SARS-CoV-2 seroprevalence and COVID-19 cases in the DRC official reports [22]. The low morbidity in our study contrasts somewhat with provincial data which recorded 603 COVID-19 related deaths since in the province since March 2020, making North-Kivu province the second worst COVID-19 affected province after Kinshasa. One plausible explanation is that vulnerable people such as the elderly and immunocompromised individuals were the most severely affected by COVID-19, and this probably resulted in most COVID-19-related case fatalities. Therefore, if the current national COVID-19 vaccination rate (below 3%) can be used as an indicator of the weakness of the vaccination campaign in the country, DRC policy makers should develop a workable policy focusing on protecting the vulnerable people through targeted vaccination, rather than continuing with an ineffective vaccination campaign that has failed miserably.

In conclusion, our study found a high seroprevalence of anti-SARS-CoV-2 antibodies among market vendor and market customers in Goma, DRC. Despite a complete lack of vaccination against SARS-CoV-2, there was no significant mortality and morbidity reported in this cohort. Such a low health impact cannot be attributed to the country’s official policy for combatting the COVID-19 pandemic but rather to the population’s enhanced capacity for acquired immunity to infectious diseases, including COVID-19, and to its youth. Our results confirm and strengthen those obtained in the same period of time in a slum of Bukavu [6] and question the relevance of vaccination in these communities. However, data from the countryside are necessary to draw up a more thorough epidemiological map of the COVID-19 in the DRC and to assess the potential health real impact on other vulnerable populations.

## Data Availability

The databases used and/or analyzed during the current study are available from the corresponding author upon request.

## Acknowledgments

We appreciate the market executives and the outreach team’s assistance in raising public awareness of the study and assisting participants with the questionnaire.

## Disclaimer

The findings and conclusions in this study are those of the authors and do not necessarily represent the official position of their respective institutions

## Ethics approval statement

This study was reviewed and approved by the Ethical Review Committee of the Université Catholique de Bukavu (number UCB/CIES/NC/02312021). All participants or their guardians (in the case of children) provided written consent before enrolment.**Table 1** Socio-demographic characteristics and SARS-CoV-2 serology of vendors and customers in two Goma markets

